# A multi-scale integrated analysis identifies KRT8 as a pan-cancer early biomarker

**DOI:** 10.1101/2020.10.01.20205450

**Authors:** Madeleine K. D. Scott, Michael G. Ozawa, Pauline Chu, Maneesha Limaye, Viswam S. Nair, Steven Schaffert, Albert C. Koong, Robert West, Purvesh Khatri

## Abstract

An early biomarker would transform our ability to screen and treat patients with cancer. The large amount of multi-scale molecular data in public repositories from various cancers provide unprecedented opportunities to find such a biomarker. However, despite identification of numerous molecular biomarkers using these public data, fewer than 1% have proven robust enough to translate into clinical practice^1^. One of the most important factors affecting the successful translation to clinical practice is lack of real-world patient population heterogeneity in the discovery process. Almost all biomarker studies analyze only a single cohort of patients with the same cancer using a single modality. Recent studies in other diseases have demonstrated the advantage of leveraging biological and technical heterogeneity across multiple independent cohorts to identify robust disease biomarkers. Here we analyzed 17149 samples from patients with one of 23 cancers that were profiled using either DNA methylation, bulk and single-cell gene expression, or protein expression in tumor and serum. First, we analyzed DNA methylation profiles of 9855 samples across 23 cancers from The Cancer Genome Atlas (TCGA). We then examined the gene expression profile of the most significantly hypomethylated gene, *KRT8*, in 6781 samples from 57 independent microarray datasets from NCBI GEO. *KRT8* was significantly over-expressed across cancers except colon cancer (summary effect size=1.05; p < 0.0001). Further, single-cell RNAseq analysis of 7447 single cells from lung tumors showed that genes that significantly correlated with *KRT8* (p < 0.05) were involved in p53-related pathways. Immunohistochemistry in tumor biopsies from 294 patients with lung cancer showed that high protein expression of KRT8 is a prognostic marker of poor survival (HR = 1.73, p = 0.01). Finally, detectable KRT8 in serum as measured by ELISA distinguished patients with pancreatic cancer from healthy controls with an AUROC=0.94. In summary, our analysis demonstrates that KRT8 is (1) differentially expressed in several cancers across all molecular modalities and (2) may be useful as a biomarker to identify patients that should be further tested for cancer.

## Introduction

Most of the public health burden of cancer results from our inability to detect tumors before they become untreatable^2^. For instance, non-small cell lung cancer (NSCLC), the leading cause of cancer deaths worldwide, progresses from early to advanced stages over a year^3^. Early detection of NSCLC is shown to substantially improve survival through surgical resection of the tumor^4^; however, after the cancer has metastasized, surgical intervention does not improve patient outcomes^5^. This critical need for early cancer biomarkers motivated the creation of consortiums like the TCGA^6^. Since the first TCGA data was released in 2006, there have been hundreds of putative molecular biomarkers proposed across all cancer types, with most focusing on gene expression biomarkers^7,8^. However, most gene signature biomarkers were identified in only one cancer type or subtype, and very few ever proved to be viable for clinical use^8,9^. Many proposed signatures failed to translate into clinical practice because they could not be replicated in outside cohorts or performed poorly when clinical data was considered^10^.

DNA methylation profiles have been shown to carry additional information to either genomic or expression data^11,12^. Yang *et al*. demonstrated that TCGA methylation data could identify clinically relevant subsets of patients with breast cancer that could not be classified by gene expression^13^. Others have documented the prognostic ability of other epigenetic signatures in colon, lung, and pancreatic cancer^14-16^.

However, the bulk of putative methylation biomarkers are limited to a single disease and face the same clinical translation issues as gene expression biomarkers^17^. To increase the probability that a methylation biomarker is useful in clinical practice, it is critical to demonstrate a robust functional and translational relevance of the differentially methylated genes in multiple cohorts^18^. Additionally, the focus on single-cancer biomarkers has raised concerns about the potential to overlook common epigenetic drivers of cancer^19^.

In this study, we performed a pan-cancer analysis of TCGA DNA methylation data from 9855 tissue samples across 23 cancers to inform subsequent gene expression, proteomic, and clinical outcome analyses. The methylation samples were divided into discovery (2019 samples across 10 cancers) and validation (7836 samples across 21 cancers). *KRT8* was the most significant differentially methylated gene across cancers. We next examined the gene expression profile of *KRT8* in 6781 samples from 57 independent microarray datasets in five solid tumor cancers (breast, colon, pancreatic, ovarian and lung) from NCBI GEO^20^, and found *KRT8* to be universally overexpressed. Our analysis of intra-cellular gene-*KRT8* expression correlations in 7447 single cells derived from lung tumor biopsies found *KRT8* is correlated with genes involved in p53-related pathways. We validated these correlations in gene expression microarrays of 1276 tissue biopsies from patients with lung cancer. We examined the prognostic relevance of tumor *KRT8* protein in 294 tissue microarrays (TMAs) from patients with lung cancer. We then calculated the prognostic value of tumor *KRT8* gene expression in pancreatic cancer with data from Protein Atlas. Finally, we validated the potential of *KRT8* as non-invasive biomarker with serum KRT8 in 32 pancreatic patients and 6 healthy controls from Stanford Hospital. An overview of this analysis is displayed in **Figure 1**.

**Figure 1.**
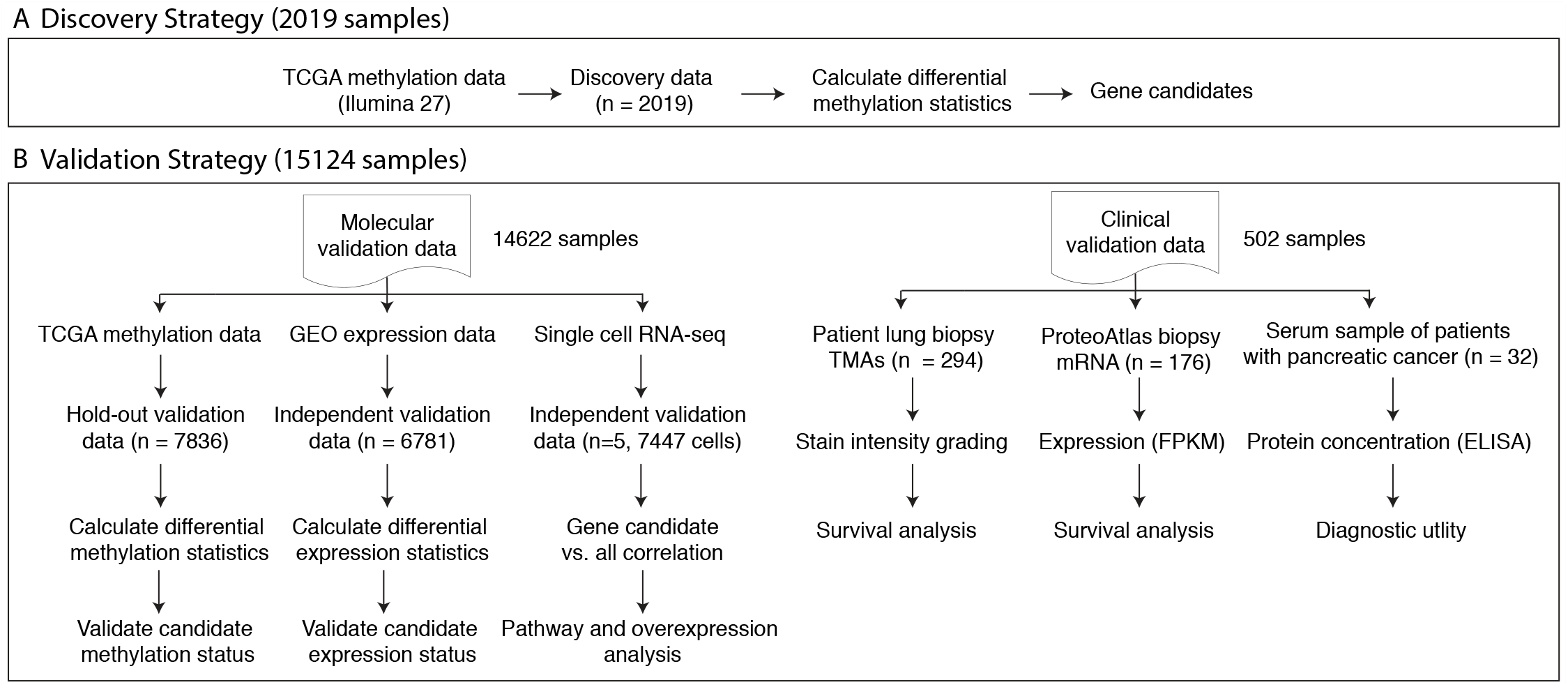
Analysis overview. **(A)** DNA methylation data from TCGA for 10 cancers comprising 2019 samples profiled using the Illumina 27 platform were used for discovery. **(B)** Validation data comprised of 15,124 samples profiled using either DNA methylation, bulk and single cell gene expression, or tumor and serum protein.

## Methods

### Data Collection from Public Repositories – TCGA and GEO

All methylation and transcriptome data used in our analyses are publicly available. We downloaded all available DNA methylation data the TCGA data portal (https://tcga-data.nci.nih.gov/tcga/tcgaHome2.jsp) irrespective of cancer on May 19, 2018. We excluded data for cancers where less than two non-cancerous samples were profiled, which resulted in DNA methylation data for 9855 samples across 23 cancers. For DNA methylation profiling, samples from these 23 cancers were profiled using either the Infinium HM27 array (27,578 CpG site targeting probes) or Infinium HM450 array (485,577 CpG site targeting probes). All data was generated and processed by The Cancer Genome Atlas research network as described previously^6,19^. We used data profiled on the HM27 array as our discovery cohort (10 cancers, 2019 samples) and data profiled on the HM450 array (21 cancers, 7836 samples) as validation.

For gene expression, we downloaded whole transcriptome data for 6,781 tumor biopsies across 57 independent datasets profiled using microarrays from the NCBI GEO. All datasets were required to measure gene expression in a minimum of two non-cancerous tissue samples. These tumor biopsies came from a patient with breast, lung, pancreatic, ovarian or colon cancer.

### Data Processing and Effect Size Estimation

We ensured all downloaded gene expression data was log2-transformed. For each gene, we calculated change in expression in a tumor biopsy as Hedges’ *g* with adjustment for small sample size because it captures both the fold change and variance. We have previously used Hedges’ g to generate robust gene signatures with diagnostic and prognostic value^21,22^. We used the random-effects inverse variance meta-analysis using Dersimonian-Laird method to calculate a summary effect size (ES) across datasets for each gene^23^. We chose Dersimonian-Laird as our previous work has shown it to be a good compromise between more conservative meta-analysis methods (Sidik–Jonkman, Hedges–Olkin, empiric Bayes, restricted maximum likelihood) and lenient methods (Hunter– Schmidt)^23^. If multiple probes mapped to a gene, the effect size for each gene was summarized via the fixed effect inverse-variance model. We corrected p-values for summary effect-sizes for multiple hypotheses testing using Benjamini-Hochberg false discovery rate (FDR) correction.^24^ We removed one cancer at a time and applied both meta-analysis methods at each iteration to avoid influence of a specific cancer with a large sample size on the results.

### Survival Analysis and Modeling

We used a right-censored model to fit survival data with the survival package in the R statistical computing environment (Version 3.5.1). We fit univariate and multivariate Cox proportional hazards models onto survival data using the coxph function. We confirmed the proportional hazard assumption with the cox.zph function.

### Human Plasma Samples

Our study includes 32 human EDTA blood plasma samples collected between January 2007 and October 2011 from identically staged patients with advanced pancreatic ductal adenocarcinoma treated at Stanford University Medical Center under an institutional review board-approved protocol. All plasma samples were collected from untreated (*de novo*) patients with biopsy-proven pancreatic adenocarcinomas. Median age at blood collection was 68 years (range 37-84 years). All patients were treated with gemcitabine-based chemotherapy and the majority also received radiotherapy. As a control group, 6 additional plasma samples were collected from age-matched, healthy volunteers under an IRB-approved protocol. Immediately after acquisition, blood samples were centrifuged and aliquots of plasma stored at -80°C.

### Enzyme-linked immunosorbent assay (ELISA)

The serum biomarker concentration was measured with a commercially available human protein sandwich enzyme immunoassay kit with two mouse monoclonal antihuman antibodies (R&D Systems, Inc., Minneapolis, MN, USA). All serum samples from patients and standards were incubated in microplate wells coated with the first mouse monoclonal anti-human biomarker antibody. After washing, a second antihuman biomarker antibody labeled with peroxidase (HRP) was added for subsequent incubation. The reaction between HRP and substrate (hydrogen peroxide and tetramethylbenzidine) resulted in color development and the intensities were measured with a microplate reader at an absorbance of 450 nm. Concentrations of serum biomarkers were determined against a standard curve.

### Single cell data collection and processing

We downloaded count matrices of 52,698 single cells from the tumor microenvironment of five lung cancer patient samples from Array Express (E-MTAB-6149)^25^. Of the total 52,698 cells, 7,447 originated from the tumor. We calculated the Pearson correlation between expression of *KRT8* and all other measured genes within each tumor cell. For each KRT8-gene correlation, we required non-zero expression of both genes in a minimum of 25 cells. We removed correlations with a p-value ≥ 0.05.

### KRT8 Expression in Patients with Pancreatic Cancer from The Protein Atlas

We downloaded prognostic information for 176 pancreatic cancer patients stratified by tumor KRT8 expression from The Protein Atlas^26^ (https://www.proteinatlas.org/ENSG00000170421-KRT8/pathology/tissue/pancreatic+cancer). We stratified patients based on median KRT8 expression of the cohort. Patient samples originated from the TCGA data repository. All counts are reported as Fragments Per Kilobase of exon per Million reads (FPKM).

### TMA cohort, and immunohistochemistry

Patient samples were retrieved from the surgical pathology archives at the Stanford Department of Pathology and linked to a clinical database using the Cancer Center Database and STRIDE Database tools from Stanford. Patients who had surgically treated disease and paraffin embedded samples from 1995 through June, 2010 were included. Surgical specimens that contained viable tumor from slides were reviewed by a board-certified pathologist (RBW) to build the Stanford Lung Cancer TMA as described previously. The area of highest tumor content was marked for coring blocks corresponding to the slides using 0.6 mm cores in duplicate arrays as previously described^27^. These cores were aligned by histology and stage and negative controls included a variety of benign and malignant tissues that included normal non-lung tissue, abnormal non-lung tissue, placental markers, and normal lung^27^. Normal lung consisted of a specimen adjacent, but distinct, from tumor over the years 1995 through 2010 to assess the variability of staining by year. OligoDT analysis was performed on the finished array to assess the architecture of selected cores and adequacy of tissue content prior to target immunohistochemistry (IHC) analysis. Serial 4 µm sections were cut from FFPE specimens and processed for IHC using the Ventana BenchMark XT automated immunostaining platform (Ventana Medical Systems/Roche, Tucson, AZ). Rabbit monoclonal anti-Cytokeratin 8 (phospho S431) antibody was obtained from Abcam (ab109452, Burlingame, CA). Mouse monocolonal Anti-Cytokeratin 8 antibody was also obtained from Abcam (ab9023, Burlingame, CA). The intensity of KRT8 immunostaining was graded from 1-4 as determined by an independent pathologist who was blinded to patient outcome.

## Results

### Integrated analysis of TCGA methylation data identifies KRT8 as hypomethylated across cancers

We identified 23 cancers that had methylation data and at least two healthy controls per cancer from TCGA. We split the resulting 9855 samples into discovery cohorts (2019 samples from 10 cancers profiled using the Illumina 27 platform) and the validation cohorts (7836 samples from 21 cancers profiled using the Illumina 450 platform) for validation. In order to avoid the potential influence of a single cancer on the results due to unequal sample sizes or other unknown confounding factors among cohorts, we performed a ‘‘leave-one-cancer-out’’ analysis. We hypothesized that the resulting set of methylation sites, irrespective of the set of cancers analyzed, would constitute a robust methylation signature across cancers. We identified 1,801 differentially methylated genes (1,081 hyper- and 720 hypomethylated, FDR < 5%) across all cancers (**Figure 1A and Supplementary Figure 1A**). We did not remove differentially methylated sites with significant heterogeneity for two reasons. First, heterogeneity is expected due to known heterogeneity within and between cancers. Second, we have previously shown that when combining across multiple datasets, filtering by heterogeneity removes higher proportion of true positives than false positives^23^. In the validation cohorts, which used Illumina 450 platform, we found 1083 out of 1,801 sites were differentially methylated across all cancers (FDR < 5%; **Figure 1B** and **Supplementary Figure 1B**).

Our discovery analysis found several previously reported differentially methylated genes. The hypomethylated genes across all cancers in the discovery cohort included *CLDN4*^*28*^ (discovery ES = -1.86, p = 8.0e-7; validation ES = -0.56, p = 1.55e-06) and *SFN*^*29*^ (discovery ES = -0.96, p = 2.01e-7; validation ES = -0.94, p = 9.4e-10) that have been previously shown to promote cancer cell proliferation (Ehrlich 2009), whereas the hypermethylated genes included known tumor suppressors such as *SOX1*^*30*^ (discovery ES = 1.05, p = 3.4e-08; validation ES = 1.08, p = 4.4e-22), *TWIST*^*31*^ (discovery ES = 0.89, p = 1.5e-5; validation ES = 0.59, p = 4.3e-16), and *GATA4*^*32*^ (discovery ES = 0.92, p = 1.7e-6; validation ES = 0.38, p = 3.77e-12). *KRT8* was the most statistically significant hypomethylated gene after multiple hypothesis correction (discovery ES = -1.71, p = 3.2e-7, FDR=9.15e-6; **Figure 2A**), but was unchanged in renal clear cell carcinoma. *KRT8* was also hypomethylated in the validation cohorts across all cancers except pheochromatoma/paraganglioma and melanoma (validation ES=-0.69, p = 3.3e-15, FDR = 4.0e-14; **Figure 2B**) (**Figure 2**).

**Figure 2.**
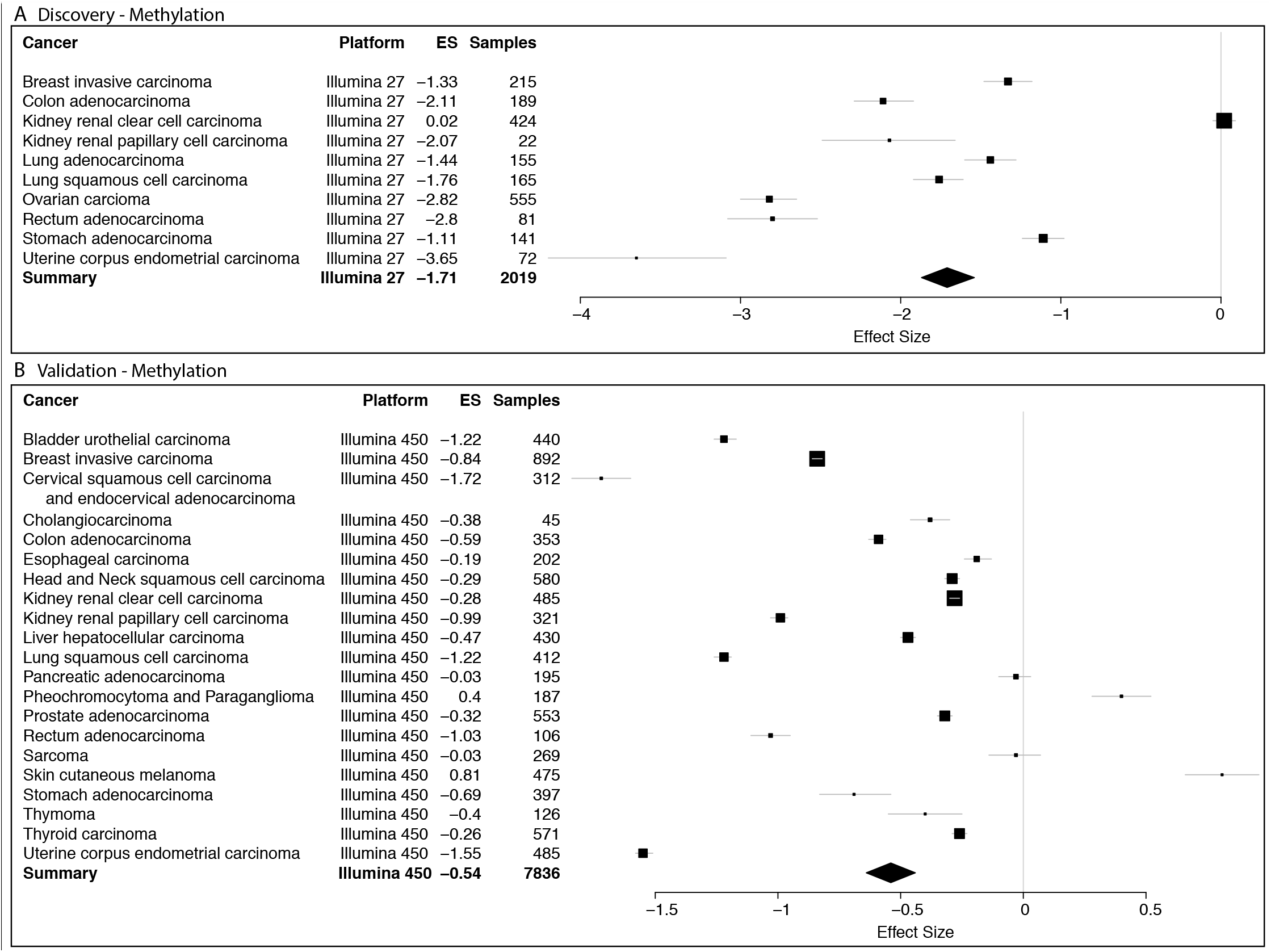
*KRT8* is hypomethylated across 23 cancers. Differential DNA methylation of *KRT8* in **(A)** discovery (10 cancers, 2019 samples, Illumina 27 platform) and **(B)** validation (21 cancers, 7836 samples, Illumina 450 platform) tumor biopsy samples compared to control non-cancerous tissue. X axes represent standardized mean difference in DNA methylation between cancer and control samples, computed as Hedges’ *g*, in log2 scale. The size of a rectangle is inversely proportional to the SEM in the corresponding cancer cohort. Whiskers represent the 95% confidence interval. A diamond represents a summary effect size for *KRT8* across cancers. Width of a diamond represents the 95% confidence interval of summary effect size. SEM: standard error of mean.

### Multi-cohort gene expression analysis demonstrates KRT8 is over-expressed in five cancers

Hypomethylation and hypermethylation typically lead to over- and under-expression of the corresponding gene, respectively.^33^ Therefore, we hypothesized that hypo- or hyper-methylated genes across multiple cancers will be over- or under-expressed across multiple cancer compared to control samples. Arguably, we could use gene expression data for the same samples from TCGA. However, we decided to use gene expression data from completely independent cohorts from a different source to increase stringency of our analysis. Therefore, to test this hypothesis, we downloaded 57 microarray gene expression datasets from the NCBI GEO^20^ comprising of 6781 samples (4870 cases, 1911 controls) obtained from human tissue biopsies of five cancers: breast, colon, lung adenocarcinoma, ovarian, or pancreatic. These 57 datasets included broad biological and technical heterogeneity, such as treatment protocols, demographics, collection year, and microarray platforms to further increase the stringency of our analysis and identify robust signals that persist despite these potential sources of vznoise.

Differential gene expression meta-analysis across all 6781 samples identified overexpression of known oncogenes such as *ERBB2* (ES =0.51, p = 6.22e-13), *KRAS* (ES =0.43, p = 2.90e-9), *CCND1* (ES = 0.25, p = 7.34e-3), and *VEGFA* (ES = 0.42, p = 2.19e-06). Housekeeping genes did not show a change in expression between control and cancer, such as *B2M* (ES = 0.12, p = .25), *HBS1L* (ES = -0.08, p = 0.15), or *EMC7* (ES = 0.18, p = 0.09)^34,35^.

Next, we calculated the Spearman correlation between the discovery methylation ES and gene expression ES in the 1,801 differentially methylated genes as -0.21 (p=1.27e-19), which in line with previous studies^36^ that examined intra-sample methylation-expression correlation (**Supplementary Figure 2**).

Finally, we found that hypomethylation of *KRT8* led to overexpression in multiple cancers compared to healthy samples (ES=1.05, p=2.8e-27, FDR=2.0e-24; **Figure 3**). *KRT8* was over-expressed in pancreatic cancer (ES=0.69, p=4.02e-08), ovarian cancer (ES=1.61, p=1.93e-03), lung cancer (ES=1.55, p=1.95e-13), and breast cancer (ES=0.88, p=7.82e-10), but not in colon cancer (ES = 0.14, p = 0.38).

**Figure 3.**
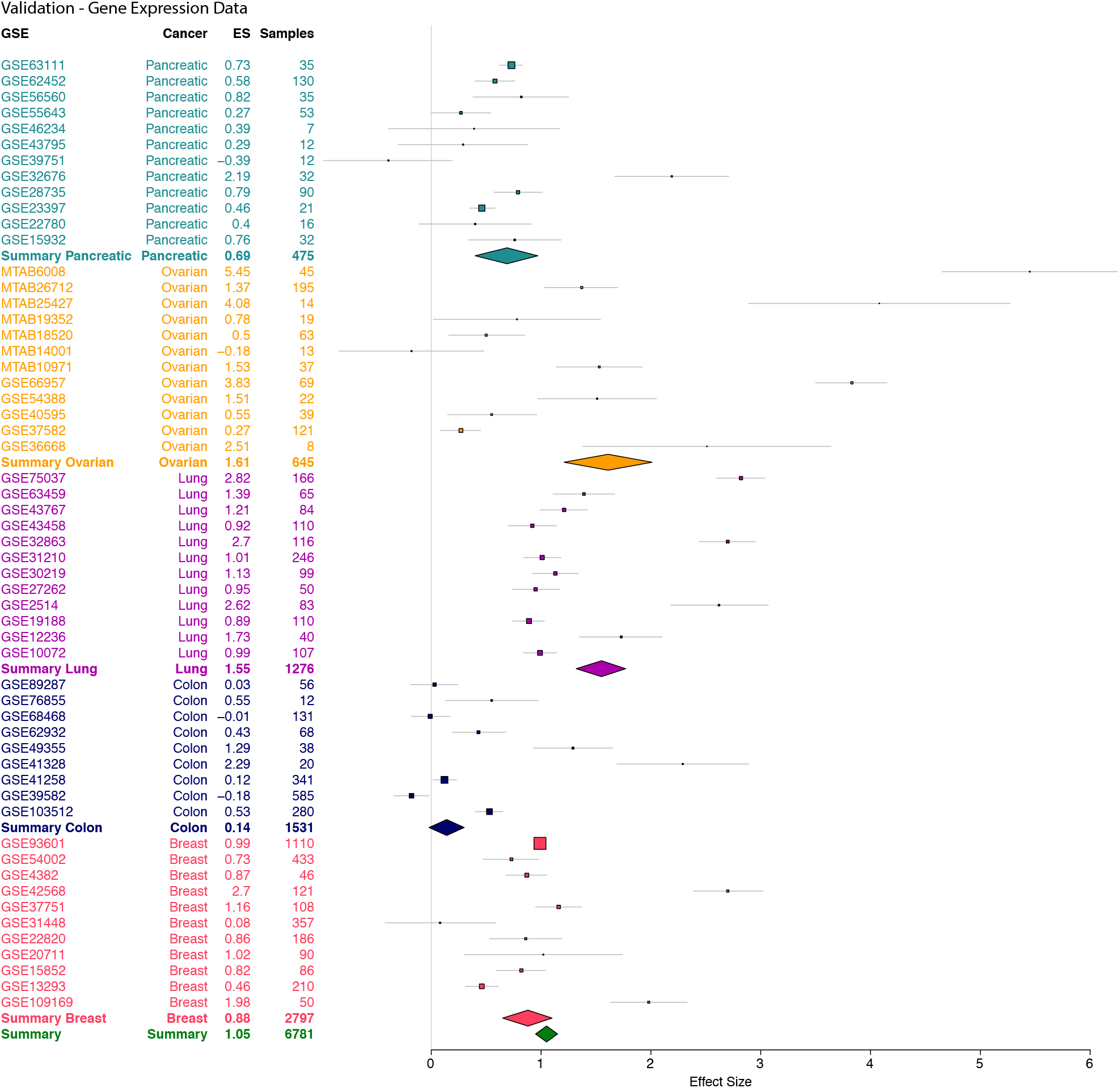
KRT8 gene is overexpressed in five cancers. The x axes represent standardized mean difference in gene expression between cancer and control samples, computed as Hedges’ g, in log2 scale. The size of a rectangle is inversely proportional to the SEM in the cohort. Whiskers represent the 95% confidence interval. A diamond represents a summary effect size for *KRT8* across cancers. Green diamond represents summary effect size across all cancers. Width of a diamond represents the 95% confidence interval of summary effect size. SEM: standard error of mean.

### KRT8 overexpression is associated with a chemotherapy-resistant phenotype in vitro

Chemotherapy resistance is responsible for more than 80% of cancer-related mortality. We investigated whether increased *KRT8* expression is associated with chemotherapy resistance. We downloaded 100 samples in seven datasets from NCBI GEO across six cancers that contained both chemotherapy-resistant and chemotherapy-sensitive cell lines. *KRT8* was consistently overexpressed across all chemo-resistant cancer cell lines (summary effect size=0.76, p=0.035; **Supplementary Figure 3**). This result demonstrates a consistent association between *KRT8* expression and chemotherapy resistance *in vitro*.

### Single cell analysis of KRT8 expression

Single cell gene expression data has allowed researchers to probe intra-cellular gene-gene correlations, which in turn suggest gene interactions or a common regulator. We analyzed intra-cellular correlations between every gene and *KRT8* with single cell RNA sequencing data of 7447 cells from tumor biopsies of five lung cancer patients. To calculate intra-cell gene-gene correlations, we correlated the expression of each gene to *KRT8* expression in every cell. Several other keratin genes were positively correlated with KRT8. For example, *KRT18* and *KRT7* had Pearson correlation of 0.59 and 0.55, respectively, with *KRT8*. Next, we preformed pathway analysis of the 100 most positively and negatively correlated genes with *KRT8* using the Reactome Knowledge Database^37^. Thirty out of the 100 genes were not annotated in the Reactome Knowledge Database. We identified six significantly enriched pathways, each of which has been previously implicated in cancer progression (**Figure 4A**). The top three significantly enriched pathways were comprised of six unique genes: *GSTP1, PRDX5, GPX2, TXNRD1, SFN, COX6A1* (**Supplementary Table 1**). Each of these six genes had an intra-cellular correlation with KRT8 expression ≥0.30 (**Figure 4A**). All genes except *GSTP1* are annotated in Reactome as involved in p53 signal transduction (**Supplementary Table 1**). However, *GSTP1* is known to be a direct transcriptional target of p53^38^, further supporting the association between *KRT8* and genes involved in the p53 pathway.

**Figure 4.**
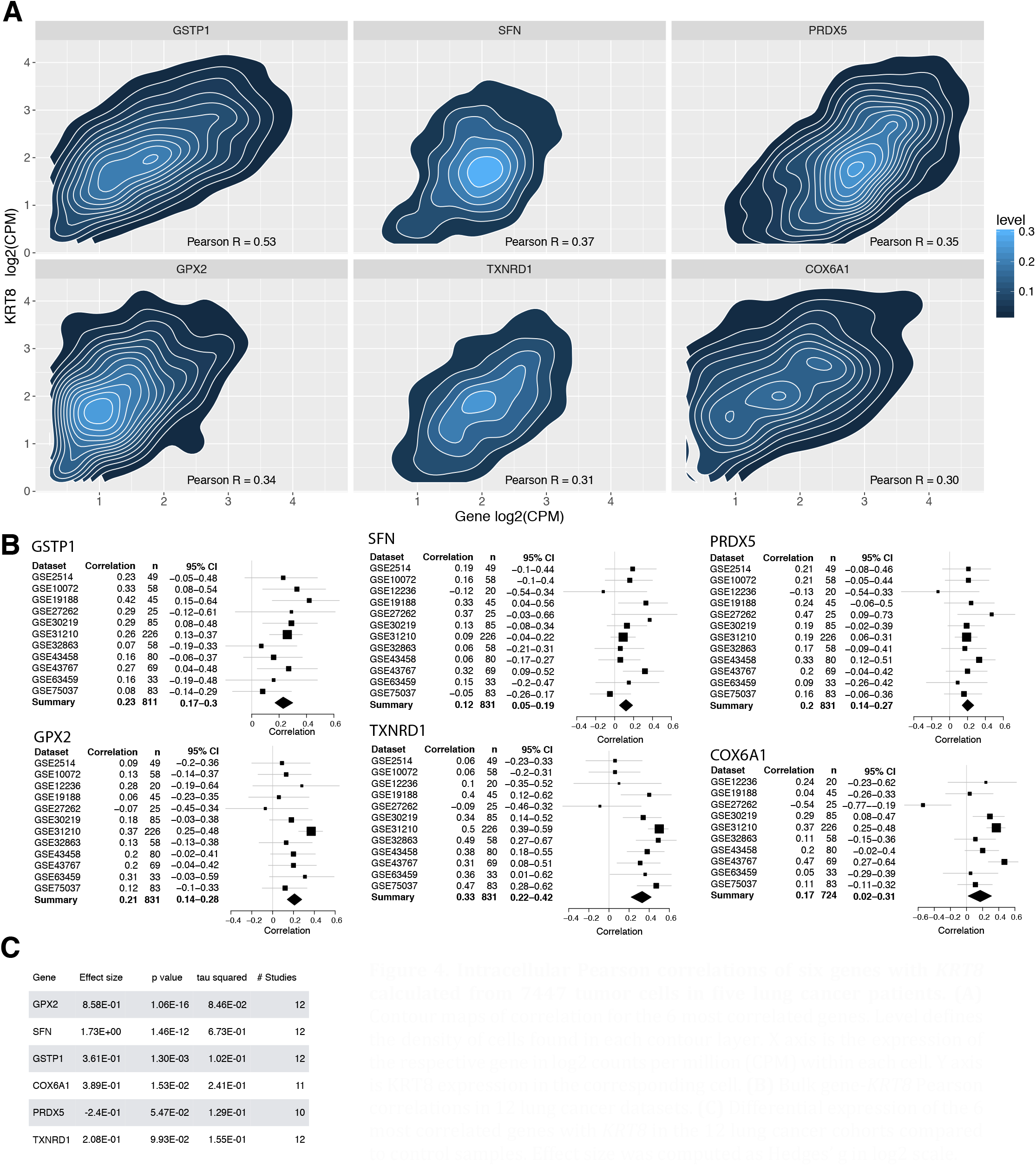
Intracellular Pearson correlations of six genes with *KRT8* calculated from 7447 tumor cells in five lung cancer patients. **(A)** Contour maps of correlation for the 6 most correlated genes. Level defines the density of cells found in each contour layer. X axis is the expression of the respective gene in log2 counts per million (CPM) within each cell. Y axis is KRT8 expression in the corresponding cell. **(B)** Bulk gene-*KRT8* Pearson correlations in 12 lung cancer datasets. **(C)** Differential expression of the 6 most correlated genes with *KRT8* in the 12 lung cancer cohorts compared to control samples. Effect size was computed as Hedges’ g in log2 scale.

We next examined the correlation between the six genes and *KRT8* in bulk lung adenocarcinoma gene expression data from microarrays of 1276 lung biopsy samples from 12 datasets. All genes were significantly correlated with *KRT8* at sample level (**Figure 4B)**. All genes except *PRDX5* were overexpressed in lung adenocarcinoma compared to healthy patients (**Figure 4C**). The majority of these six genes were additionally overexpressed across 5505 microarray samples from four cancers (breast, colon, ovarian, and pancreatic; **Supplementary Table 2**).

### Protein expression of KRT8 is associated with poor outcomes in patients with lung adenocarcinoma

Given robust hypomethylation of *KRT8* across 9,855 samples from 23 cancers, over-expression across 6,781 biopsies from 5 cancers, strong association with chemo-resistance, and sustained correlation with p53-regulated genes both at single-cell and sample levels, we investigated whether *KRT8* is also expressed at protein-level in tumor biopsies, and whether it is associated with survival in patients with either lung adenocarcinoma or lung squamous cell carcinoma. We stained tissue microarrays (TMAs) containing 294 lung tumors (228 lung adenocarcinoma, 66 lung squamous cell carcinoma) resected from patients at Stanford Hospital for *KRT8* protein (**Supplementary Table 3**). An expert pathologist (MO) rated the maximum intensity of cancerous cell *KRT8* staining in each TMA (**Figure 5A**). Out of the 294 samples, 5 (1.7 %) scored as 1+, 35 (11.9%) as 1-2+, 55 (18.7%) as 1-3+, 8 (2.72%) as 2+, 85 (28.9%) as 2-3+ and 106 (36.1%) as 3+. In a multivariable cox regression model, *KRT8* intensity was a significant predictor of mortality after adjusting for sex and age at diagnosis in lung adenocarcinoma (Hazard Ratio = 1.49, 95% CI = 1.06 – 2.10, p=0.02), but not in squamous cell (Hazard Ratio = 1.19, 95% CI = 0.57 – 2.52, p=0.65; **Figure 5B**).

**Figure 5.**
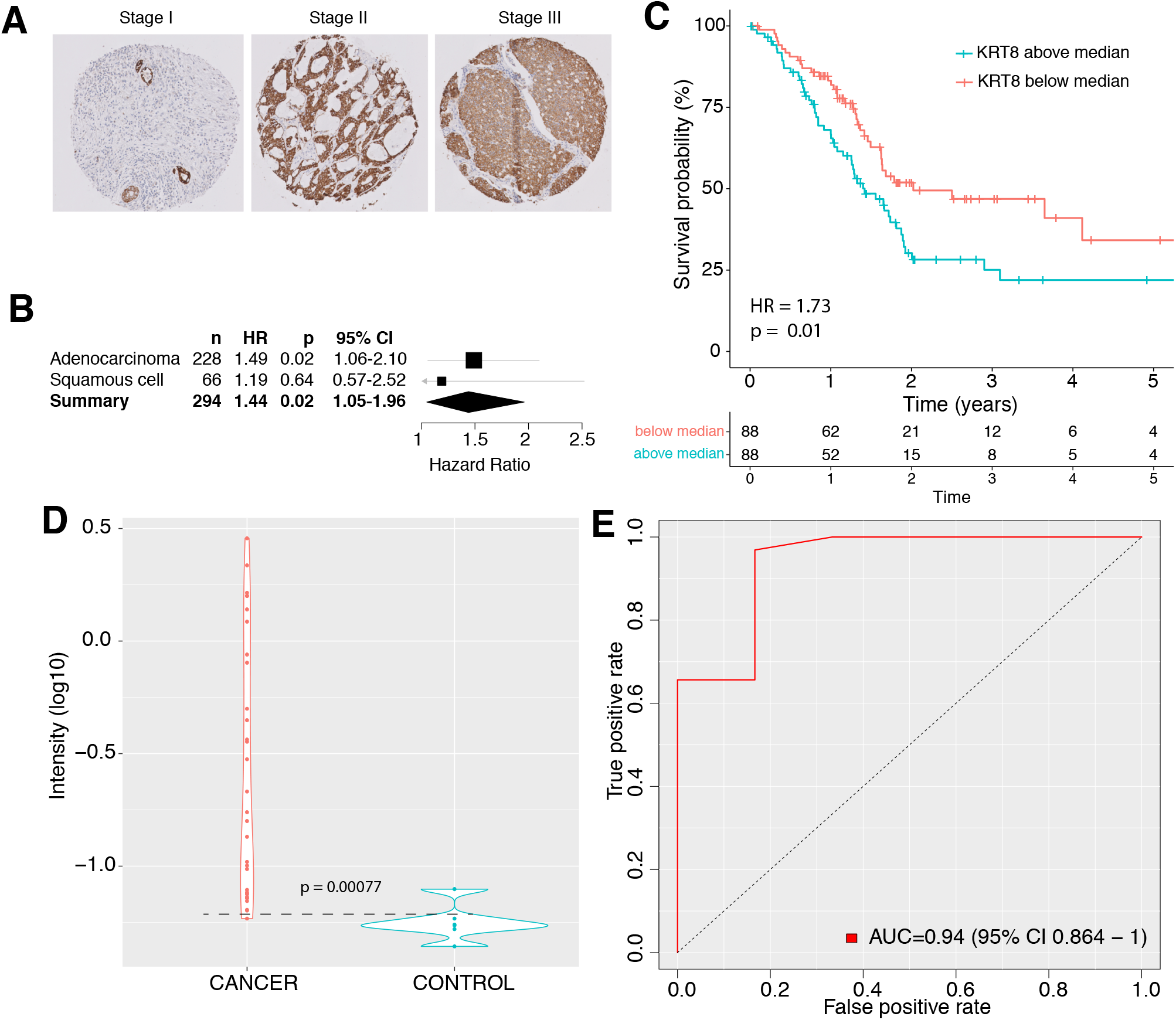
Protein measurement of KRT8 in cancer. **A**. IHC of lung adenocarcinoma TMAs for KRT8. **B**. Cox proportional hazard of 294 lung cancer samples stratified by KRT8 concentration. **C**. Survival of 176 patients with pancreatic cancer stratified by KRT8 expression relative to the median of the cohort. **D-E**. Violin (**D**) and ROC (**E**) plots of serum KRT8 as measured by ELISA in patients with pancreatic cancer and healthy controls. Dashed line represents the ELISA detection threshold. Width of a violin plot indicates density of samples, where each dot represents a sample.

### Higher RNA expression of KRT8 is associated with reduced survival in patients with pancreatic cancer

Next, we investigated whether *KRT8* tumor gene expression is a prognostic marker of survival. We downloaded *KRT8* expression and corresponding survival data for 176 patients with stage I-IV pancreatic cancer from Human Protein Atlas (**Supplementary Table 4**). We classified patients as either “High *KRT8*” or “Low *KRT8*” if their *KRT8* expression was above or below the median *KRT8* expression of the cohort (363.5 FPKM), respectively. Patients in the “High *KRT8*” group had an increased risk of mortality (cox proportional hazard ratio = 1.73 p = 0.01 **Figure 5C**).

### Serum KRT8 discriminates between healthy and pancreatic patients and correlates with survival time

Finally, we explored the potential of KRT8 as a minimally invasive biomarker. We measured *KRT8* concentration in serum of 32 biopsy-confirmed patients with pancreatic ductal adenocarcinoma and six healthy controls by enzyme-linked immunosorbent assays (ELISA). Samples were collected from Stanford Hospital (**Supplementary Table 5**). The mean KRT8 concentration was significantly higher in the pancreatic cancer patients compared to that of healthy controls (p = 7.7e-4; **Figure 5D**). Samples were considered KRT8+ if they had a measured KRT8 value about the detectability limit of the ELISA (0.06 RLU). KRT8+ status distinguished patients with pancreatic cancer from healthy controls with an area under the curve (AUC) of 0.94 (**Figure 5E**) and an area under the precision recall curve (AUPRC) of 0.99 (**Supplementary Figure 4**).

## Discussion

Only a fraction of molecular cancer biomarkers published in academic literature are reproducible in follow-up studies. The first step to identifying a robust biomarker is to ensure that the discovery phase has included a heterogeneous set of samples, platforms, and measurement technologies. Here, we identified KRT8 as such a biomarker by integrating DNA methylation profiling of 2019 samples across 10 cancers from the TCGA. We then validated that KRT8 is a robust biomarker on 7836 samples in 21 cancers measured with a different DNA methylation platform within the TCGA. We next analyzed the diagnostic and prognostic value of tumor *KRT8* gene and protein expression as well as serum KRT8 using ELISA in over 7000 samples spanning 10 years, multiple platforms, and data repositories.

Pan-cancer methylation findings have been hindered by questions about batch effects and platform bias^39^. In this work, we used samples run on Illumina 27 platform as our discovery data and Illumina 450 as validation. KRT8 was significantly hypomethlyated in both platforms, suggesting it is robust to platform bias. While TCGA has gene expression data, we chose to use microarray samples from the NCBI GEO to ensure that our findings would be robust to data type, batch effect, and platform.

Single cell analysis has broadened our understanding of tumor heterogeneity, but it can be difficult to interpret the immediate translational value of a single time point scRNA-seq analysis. Here, we show that intra-cellular gene-gene correlations can suggest overlooked gene functions. Additionally, by replicating the correlations found at the single cell level in bulk tissue microarrays, we propose a strategy for validating expression patterns seen in the single cell level.

Our study has several limitations. First, it does not include the entirety of all cancer data available in the public sphere, and thus presents an incomplete picture of *KRT8* across all data. However, this study used 17149 samples across 23 cancers, which still includes significant amount of biological, clinical, and technical heterogeneity in the real world patient population. Further, we have previously shown that 4-5 independent datasets with a total of approximately 200-250 samples substantially increases the probability of validation in independent cohrots^23^. Second, we only required two control samples in the methylation discovery analysis, which could have led to false positive or patient-specific effects within a datasets. However, the integration of all the discovery cohorts and independent validation using Illumina 450 methylation platform substantially mitigated the effect of a single cancer outlier. In addition, our rigorous downstream analysis of gene expression from 6781 samples in 57 datasets from 5 cancers provide strong evidence of the robustness of our analyses. Third, we chose only the top gene and validated it here. It is possible that other genes may provide equal or greater prognostic value than *KRT8*. However, our aim is to demonstrate the value of the framework we propose here and thus we explored only the most promising gene, *KRT8*. Forth, we do not provide any indication of the mechanism underlying the prognostic value of *KRT8*. It may be as straightforward as increasing epithelial cancer cell numbers results in more *KRT8* released into the bloodstream, or perhaps there is a more complex biological phenomenon at work. These questions can only be answered with follow-up hypothesis-driven research.

Previous reports have identified role of *KRT8* in the progression of lung and renal cancer.^40,41^ However, *KRT8* has never been shown to be overrepresented across cancers in a multi-omic analysis. One GEO dataset (GSE15932) contained expression from peripheral blood samples. In this dataset, *KRT8* expression distinguished cancerous from healthy patients, suggesting that circulating *KRT8* RNA may be a candidate for a diagnostic blood biomarker. Biomarkers not only have diagnostic and prognostic implications, but are also helpful for measurement of treatment responses, surveillance for tumor recurrence and guiding clinical decisions. For many cancers, there is not a single blood biomarker; others like pancreatic cancer have one or two unreliable screening biomarkers. CA19-9 is used as a biomarker in pancreatic cancer, but due to its limitations and the low prevalence of pancreatic cancer is only used to monitor for reoccurrence.^42^ Here we show the potential use of serum KRT8 protein as a blood biomarker in pancreatic cancer. Given that we identified *KRT8* as overexpressed across cancers, it stands to reason that KRT8 may be useful as a peripheral biomarker in other cancers as well.

Most importantly, this work demonstrates a strategy to translate large molecular analyses into specific, clinically relevant hypotheses. Omics sciences enable complex biological systems to be visualized in a holistic and integrative manner. Application of systems biology to interpret large multidimensional omics data across cancer types will enable the robust identification of biomarkers that share common pathophysiology, which can potentially be further explored for pan-cancer interventions

## Supporting information

All Other Supplement

## Data Availability

Microarray data are available from the NCBI GEO at: https://www.ncbi.nlm.nih.gov/geo/. The accession numbers and corresponding links for the individual studies are listed in Supplemental Table 6.

## Ethics approval and consent to participate

All aspects of this study were approved by the Stanford Institutional Review Board in accordance with the Declaration of Helsinki guidelines for the ethical conduct of research. The reference number for the approval is IRB-20170. A waiver of informed consent was obtained for the subjects in this study according to Stanford’s Institutional Review Board policy since this was a retrospective study of both alive and deceased patients, many of whom were lost to follow-up.

## Availability of data and materials

Microarray data are available from the NCBI GEO at: https://www.ncbi.nlm.nih.gov/geo/ The accession numbers and corresponding links for the individual studies are listed in Supplemental Table 6.

## Acknowledgements

We would like to acknowledge Julien Sage for his constructive feedback throughout this project.

## Notes

### Competing Interest Statement

The authors have declared no competing interest.

### Funding Statement

This work was supported in part by the Bill and Melinda Gates Foundation, and grants RO1 AI125197-01, U19AI109662, and U19AI057229 from the National Institute for Allergy and Infectious Diseases to P.K.; grant F30 HL149252-01A1 from the National Heart, Lung, and Blood Institute to M.K.D.S.

