## Supplementary material for "A multi-scale integrated analysis identifies KRT8 as a pan-cancer early biomarker": All Other Supplement

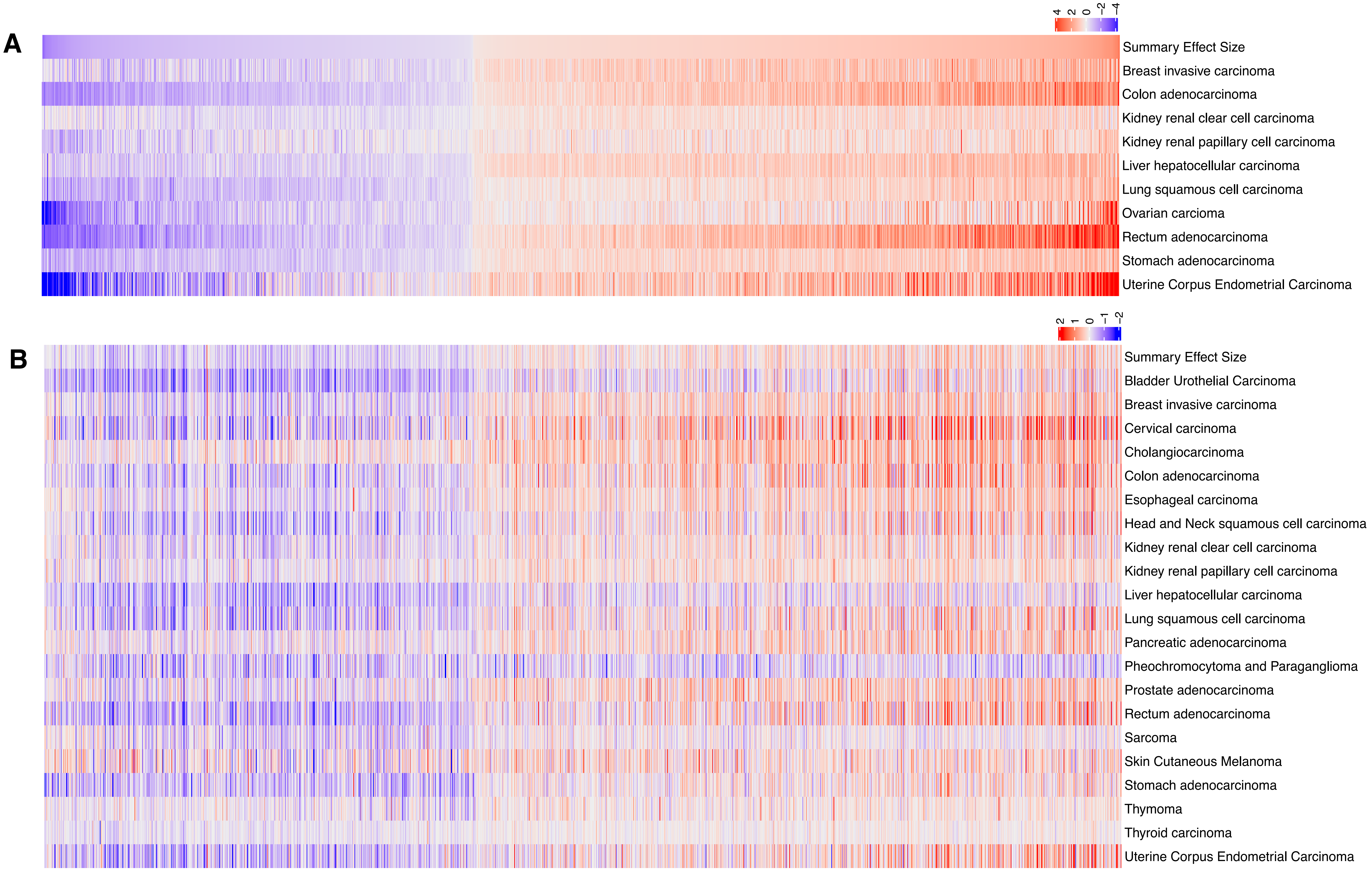

**Supplemental Figure 1**. 1,801 differentially methylated genes (1,081 hyper- and 720 hypomethylated, FDR < 5%) across all cancers. Top row indicates the summary effect size across all cancers in **(A)** the discovery data using the Illumina 27 platform and **(B)** validation data on the Illumina 450 platform. Gene order is maintained between **(A)** and **(B)**.

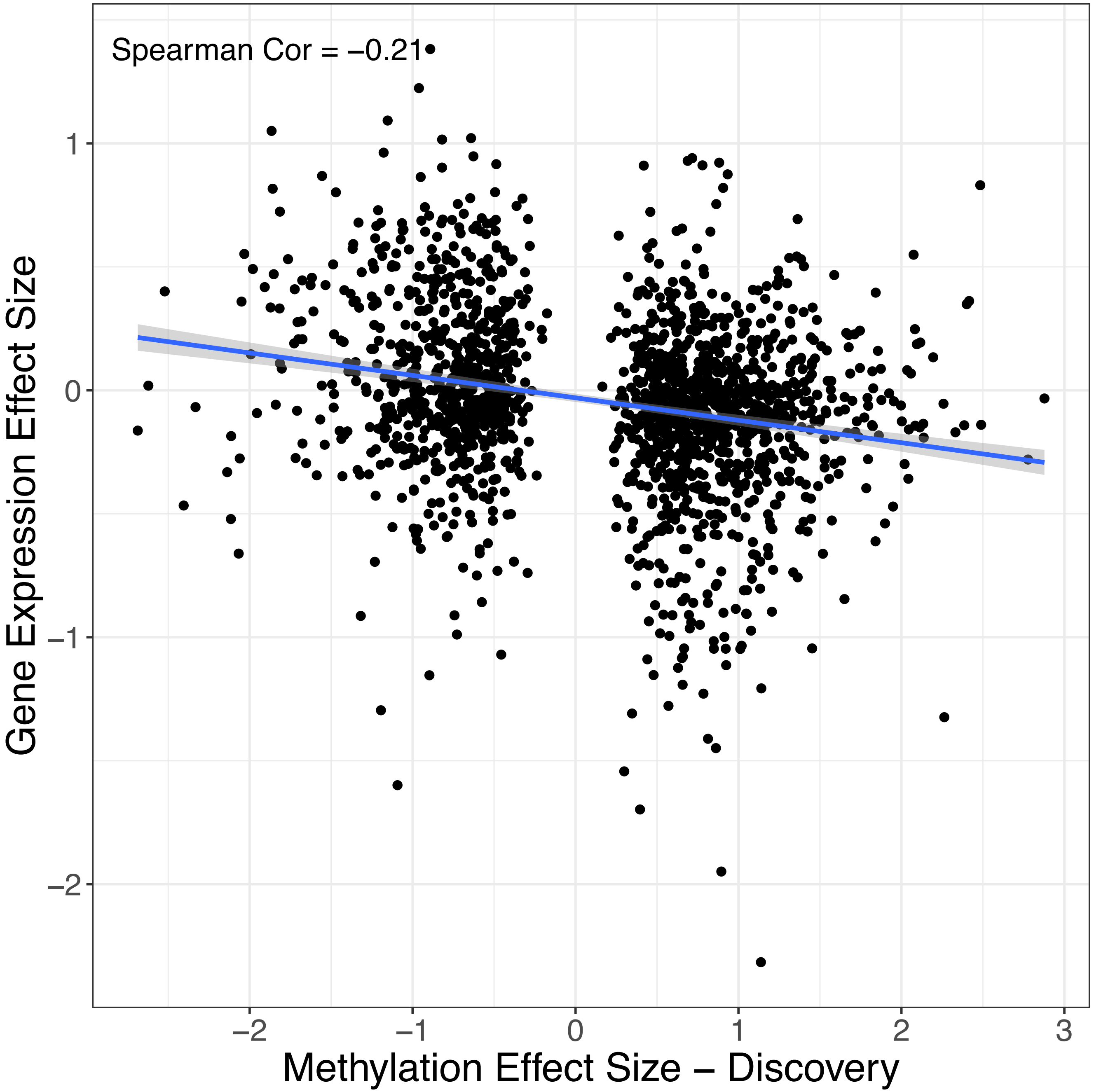

**Supplemental Figure 2.** Spearman correlation between the discovery methylation effect size and gene expression effectsize across the 1,801 differentially methylated genes

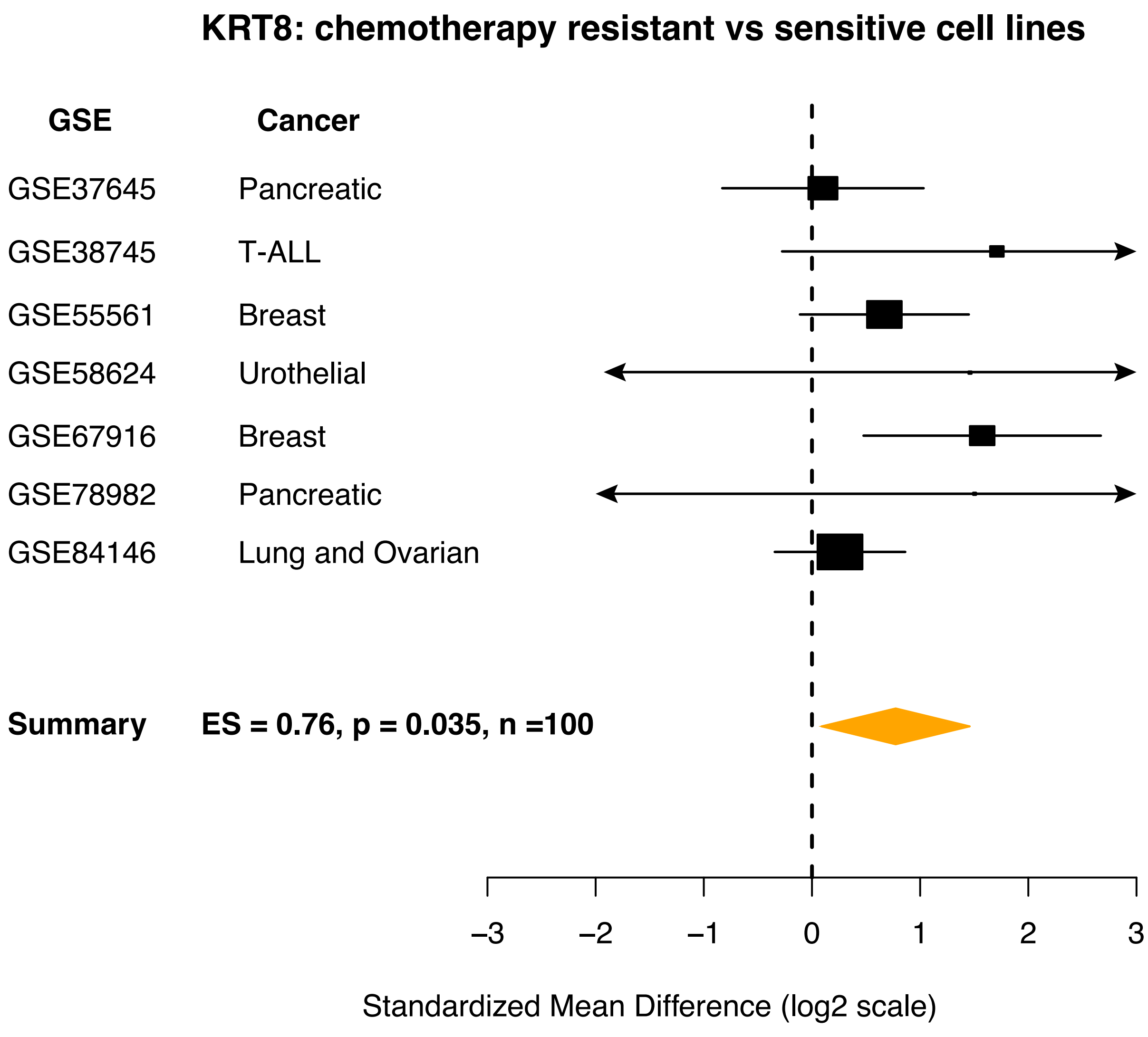

**Supplemental Figure 3.** Expression of KRT8 in chemotherapy resistant cancer cell lines compared to chemotherapy sensitive cell lines in the same tissue across seven independent datasets. ES = Effect size.

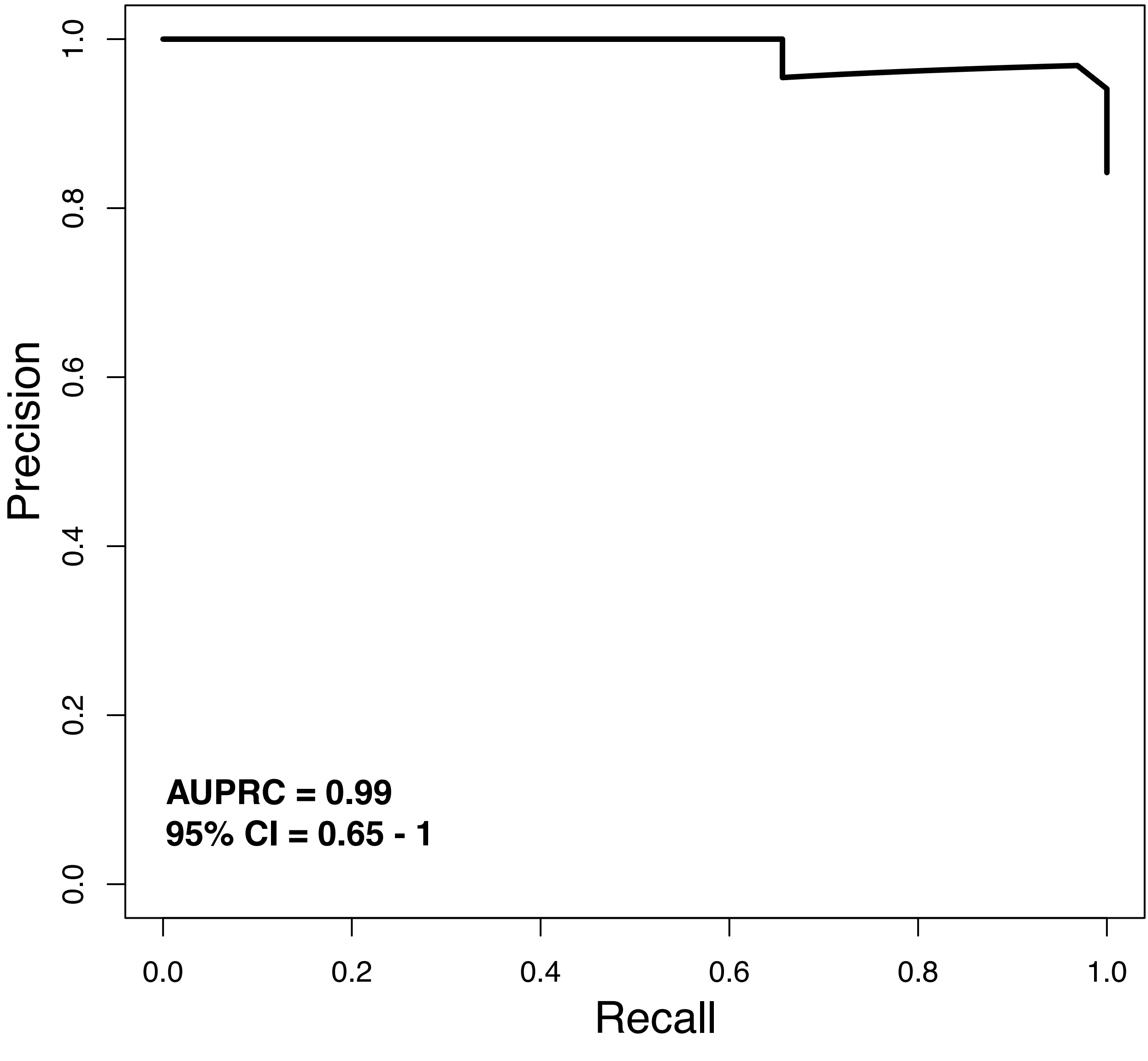

**Supplemental Figure 4**. Precision-Recall curve for desciminating pancreatic cancer patients (n = 32) from healthy controls (n = 6) by serum KRT8 as measured by ELISA.

**Supplemental Table 1.** Reactome pathway analysis of 100 most KRT8-correlated and anticorrelated genes. Correlations were determined from expression levels within single cells.

| Upregulated | Pathway | p.Geomean | Stat mean | P val | qval | set size | exp1 |
|---|---|---|---|---|---|---|---|
| | GO:0034220 ion transmembrane transport | 0.021 | 2.185 | 0.021 | 0.921 | 15.000 | 0.021 |
| | GO:0007017 microtubule-based process | 0.030 | 1.975 | 0.030 | 0.921 | 15.000 | 0.030 |
| | GO:0006091 generation of precursor metabolites and energy | 0.039 | 1.884 | 0.039 | 0.921 | 12.000 | 0.039 |
| | GO:0000226 microtubule cytoskeleton organization | 0.040 | 1.866 | 0.040 | 0.921 | 10.000 | 0.040 |
| | GO:0045088 regulation of innate immune response | 0.062 | 1.633 | 0.062 | 0.921 | 12.000 | 0.062 |
| | GO:0034097 response to cytokine stimulus | 0.064 | 1.544 | 0.064 | 0.921 | 31.000 | 0.064 |
| Down | | | | | | | |
| | GO:0007155 cell adhesion | 0.006 | -2.554 | 0.006 | 0.822 | 49.000 | 0.006 |
| | GO:0022610 biological adhesion | 0.006 | -2.554 | 0.006 | 0.822 | 49.000 | 0.006 |
| | GO:0030155 regulation of cell adhesion | 0.007 | -2.674 | 0.007 | 0.822 | 14.000 | 0.007 |
| | GO:0016337 cell-cell adhesion | 0.009 | -2.497 | 0.009 | 0.822 | 19.000 | 0.009 |
| | GO:0051336 regulation of hydrolase activity | 0.027 | -1.956 | 0.027 | 0.822 | 42.000 | 0.027 |
| | GO:0006790 sulfur compound metabolic process | 0.045 | -1.810 | 0.045 | 0.822 | 10.000 | 0.045 |

**Supplementary Table 2**. Correlation of six genes with KRT8 expression in bulk tissue microarrays

| Cancer | Gene Name | effectSize | effectSizeStandardError | effectSizePval | effectSizeFDR | # Studies |
|---|---|---|---|---|---|---|
| Lung adenocarcinoma | GPX2 | 0.85825435 | 0.10343277 | 1.06E-16 | 8.33E-16 | 12 |
| | SFN | 1.73484346 | 0.24509068 | 1.46E-12 | 7.97E-12 | 12 |
| | GSTP1 | 0.36090358 | 0.11225354 | 0.0013041 | 0.0025582 | 12 |
| | COX6A1 | 0.38921725 | 0.16041534 | 0.01525326 | 0.02462262 | 11 |
| | PRDX5 | -0.2398083 | 0.12480822 | 0.05467952 | 0.07900236 | 10 |
| | TXNRD1 | 0.20785324 | 0.12609836 | 0.09928248 | 0.13484462 | 12 |
| Breast invasive carcinoma | GPX2 | -0.2375308 | 0.04801474 | 7.54E-07 | 1.35E-05 | 11 |
| | GSTP1 | -0.3034343 | 0.08920859 | 0.00067041 | 0.00381338 | 11 |
| | SFN | 0.61918602 | 0.25434585 | 0.01491545 | 0.04712083 | 9 |
| | COX6A1 | 0.34303382 | 0.19992861 | 0.08620203 | 0.19004273 | 10 |
| | PRDX5 | 0.06965973 | 0.15040264 | 0.64325327 | 0.78503691 | 10 |
| | TXNRD1 | -0.013877 | 0.09478392 | 0.88360029 | 0.937214 | 11 |
| Colon adenocarcinoma | TXNRD1 | 0.29526367 | 0.14820284 | 0.04633878 | 0.57457234 | 9 |
| | SFN | 0.16939059 | 0.14314671 | 0.23667617 | 0.81649353 | 9 |
| | COX6A1 | 0.21129821 | 0.18870124 | 0.26282031 | 0.83222568 | 10 |
| | GSTP1 | 0.11125449 | 0.18337014 | 0.54403616 | 0.93600853 | 9 |
| | GPX2 | -0.0618429 | 0.1610988 | 0.70106622 | 0.96388548 | 10 |
| | PRDX5 | -0.0106303 | 0.1627399 | 0.9479186 | 0.99373678 | 7 |
| Pancreatic carcinoma | SFN | 1.55433781 | 0.15874762 | 1.23E-22 | 5.72E-20 | 11 |
| | GSTP1 | 1.24803455 | 0.19765895 | 2.72E-10 | 1.26E-08 | 11 |
| | GPX2 | 1.09113993 | 0.17950857 | 1.21E-09 | 4.79E-08 | 12 |
| | TXNRD1 | 0.79114389 | 0.21840454 | 0.00029191 | 0.00173964 | 12 |
| | PRDX5 | 0.78212231 | 0.22006658 | 0.00037938 | 0.00214582 | 11 |
| | COX6A1 | 0.17429096 | 0.16489239 | 0.2905124 | 0.40571164 | 11 |
| Ovarian carcioma | SFN | 1.60402136 | 0.26433707 | 1.29E-09 | 1.11E-07 | 12 |
| | GSTP1 | 1.33878562 | 0.46126096 | 0.00370259 | 0.0316481 | 12 |
| | COX6A1 | 0.59523921 | 0.23225486 | 0.01038107 | 0.06663287 | 12 |
| | TXNRD1 | -0.4461765 | 0.23997164 | 0.06298629 | 0.22142444 | 12 |
| | GPX2 | -0.2316358 | 0.18909074 | 0.22057584 | 0.46257419 | 12 |
| | PRDX5 | 0.17328605 | 0.39255309 | 0.65889926 | 0.81042429 | 10 |

**Supplementary Table 3.** Demographic information for the adenocarcinoma patients profiled by tissue microarray.

| | |
|---|---|
| n | 294 |
| Grade (%) | |
| well | 39 (13.3) |
| moderate | 167 (56.8) |
| poor | 82 (27.9) |
| undifferentiated | 1 (0.3) |
| not stated | 4 (1.7) |
| Age (mean (SD)) | 67.70 (10.49) |
| Sex = male (%) | 135 (45.9) |
| Primary tumor size (mean (SD)) | 3.45 (2.00) |
| Subtype (%) | |
| Adenocarcinoma | 228 (77.6) |
| Squamous cell | 66 (22.4) |
| Stage (mean (SD)) | 1.64 (0.76) |

**Supplementary Table 4**. Demographic information for the 176 patients with pancreatic cancer.

| Characteristic | Overall |
|---|---|
| n | 176 |
| Age (mean (SD)) | 64.85 (10.77) |
| Male (%) | 96 (54.5) |
| Stage (%) | |
| i | 1 ( 0.6) |
| ia | 5 ( 2.8) |
| ib | 15 ( 8.5) |
| iia | 28 (15.9) |
| iib | 117 (66.5) |
| iii | 3 ( 1.7) |
| iv | 4 ( 2.3) |
| not reported | 3 ( 1.7) |
| Days observed (mean | 570.65 (478.36) |
| FPKM (mean (SD)) | 386.77 (218.16) |
| Died (%) | 92 (52.3) |

**Supplementary Figure 5**. Demographic information on 32 patients with pancreatic cancer.

| | |
|---|---|
| Patients' age (years), mean (range) | 69 (43 - 85) |
| Female, *n* (%) | 12 (38) |
| Tumor grade, *n* (%) | |
| Well differentiated | 7 (22) |
| Moderately differentiated | 19 (59) |
| Poorly differentiated | 6 (19) |
| Lymph node status, *n* (%) | |
| Positive | 16 (50) |
| Negative | 16 (50) |
| Resection margin, *n* (%) | |
| Positive | 10 (31) |
| Negative | 22 (69) |
| Tumor size (cm), median (range) | 3.80 (1.10-6.00) |
| Bilirubin (mg/dL), mean (range) | 0.65 (0.5-1.3) |
| CA19-9 (U/ml), mean (range) | 26059 (1-780486) |
